# Impact of Traditional Korean Medicine Use on Medical Cost in Patients with Cancer in Korea: A Cross-Sectional Analysis

**DOI:** 10.1101/2025.04.25.25326422

**Authors:** Ji-eun Yu, Hui-Yong Kwak, Eunji Ahn, Dongsu Kim

## Abstract

**Background:** Cancer is a major cause of death, and patients with cancer use traditional medicine (TM) or complementary and alternative medicine (CAM) to control cancer-related symptoms. Although previous studies have shown that TM and CAM can improve specific conditions in patients with cancer, results regarding their economic utility are inconclusive. The traditional Korean medicine (TKM) coexists with Western medicine (WM) under a formal licensing system, and some uses of CAM are not prescribed by TKM doctors. This study aimed to investigate the use of TKM in patients with cancer and its costs and to confirm the role of TKM by revealing the correlation between WM costs and unprofessional CAM costs.

**Methods:** We used the Korean Medical Panel Data from 2015 to 2018 from the Korea Institute for Health and Social Affairs. T-tests were performed on the use of TKM in patients with cancer and on WM and CAM costs. A correlation analysis was conducted between TKM, WM, and CAM costs, and a generalized linear model (GLM) analysis to control for the participants’ characteristics. Regression analysis was used to analyze the effects of TKM use on quality of life.

**Results:** In total, 321 patients with cancer were identified, including 150 users and 171 non-users of TKM. The participants used TKM for an average of 2.41±7.22 times, and the average cost was 26.27±96.04 (US$). The most common injury codes were musculoskeletal and soft tissue diseases (87.64%) and the most common treatments were acupuncture, moxibustion, and cupping (96.51%). TKM use did not significantly affect WM or CAM costs, but it improved the quality of life. Moreover, TKM costs positively correlated with CAM costs.

**Conclusion:** Although the use of TKM and its costs were not significantly related to WM costs and may seem independent of each other, it may be because the purpose of TKM use in this study was not limited to cancer treatment. In addition, the quality of life of the patients with TKM improved, suggesting that the appropriate use of WM and TKM may improve the symptoms and quality of life in patients with cancer.

## 1. Background

Cancer is one of the most common causes of premature death [1]. Recently, with the development of medicines and technologies related to treatment, the age-standardized cancer mortality rate has decreased, but the 5-year survival rate is still only 45.9% to 68.0% [2]. Fear of metastasis, uncertainty about complete remission, fear of death, and concern about recurrence are anxiety factors in patients with cancer [3,4]. In addition, pain, fatigue, and nausea increase the level of anxiety and depression in the patients; thus, the need for palliative care in various dimensions is imperative [5,6].

In these situations, patients with cancer use traditional medicine (TM) and complementary and alternative medicine (CAM) to control the symptoms, treatment side effects, and emotional health, and for direct and indirect positive effects on cancer treatment [7]. Approximately 51% of patients with cancer use one type of CAM [8]. The cost of CAM increases before and after cancer diagnosis, and survivors of cancer spend approximately 25% more in CAM than the general adult population [9,10]. However, a large proportion of patients with cancer purchasing CAM supplements without a prescription from an expert raise concerns about the interaction with ongoing cancer treatments [11]. In fact, 20–77% of the patients who use CAM do not discuss it with their medical team and obtain information about CAM from non-professional sources, such as friends or family [12,13].

In East Asian countries including Japan, China, Korea, and Taiwan, TM is prescribed by licensed specialists and managed through the national healthcare system [14]. Research has focused on whether medical practices based on East Asian TM can improve the specific conditions of patients with cancer. Consequently, it has been confirmed that acupuncture and herbal medicine can manage cancer-related symptoms and improve the side effects [15–19]. However, the economic utility of CAM for patients with cancer is discussed separately from its effectiveness, and sometimes the cost of accessing CAM exceeds its utility. Although acupuncture is effective for peripheral neuropathy after chemotherapy, the results of an economic evaluation study conducted in Hong Kong were negative [20,21]. However, economic evaluations can only answer one preset question on whether a specific medical practice is performed or compare it with other practices. Thus, it cannot estimate the influence of CAM on the entire healthcare system [22]. Therefore, studies have attempted to determine whether investing resources in TM treatment is meaningful by exploring the proportion of TM in the total medical expenses of patients with cancer and by comparing the medical expense of those who chose TM with that of those who did not [23].

In Korea, traditional Korean medicine (TKM) coexists with Western medicine (WM) under a formal licensing system. Collaboration between TKM and WM actively take place, and the costs are largely covered by the National Health Insurance Service (NHIS) [24]. As with TM and CAM worldwide, with TKM in Korea, the use of medical services from non-professional sources is a problem. For example, while 11.6% of patients with cancer use TKM practices through medical institutions, the percentage increases to 31.0% including those who use herbal extracts (such as ginseng) not prescribed by a licensed practitioner, indicating that more than half of the patients use CAM prescribed by non-specialists [25]. Contrary to the general belief that herbal therapy is safe, most accidents involving the misuse of herbal medicine occur through routes that do not involve TKM doctors [26,27]. Therefore, it is a task for the TKM community to encourage the use of TKM through institutional experts. In this context, our study distinguishes between TKM and CAM, where TKM refers to medical practices performed in TKM institutions by TKM doctors who are official medical service providers, whereas CAM refers to the use of acupuncture, moxibustion, and herbal medicine provided by qualified or non-qualified persons who are not TKM doctors through pathways other than TKM institutions.

In this study, considering the particularity of the Korean medical system, we analyzed the TKM, WM, and CAM costs borne by patients with cancer to determine the correlation between the use of TKM and other fields and to infer the role of TKM. In addition, we aimed to review whether the coverage of TKM treatment received by patients with cancer in Korea is appropriately positioned within the overall healthcare system and whether it is a complementary or an alternative option.

## 2. Methods

### 2.1. Data source

We used the Korea Health Panel Survey (KHPS) data. The KHPS comprises the data collected by the Korea Institute for Health and Social Affairs (KIHASA; a research institute under the Office of the Prime Minister, established to set a national healthcare policy) and the NHIS (the only insurer of public insurance in Korea). The KIHASA and NHIS created the KHPS in 2008 to analyze the healthcare behavior of Koreans and its impact on health and healthcare finances. KHPS data is longitudinal in a panel format, and the first panel was maintained from 2008 to 2018. A sample was selected from the 2005 Korean Population and Housing Census data using regional stratification variables and has been tracked annually since 2008 [28].

Information on healthcare utilization was collected using the participants’ recorded use of healthcare in a prescribed household accounting form, and other information on the household and household members was collected through face-to-face interviews. We used the KHPS data to analyze the medical expenditures of patients with cancer (TKM, WM, and CAM costs), because it is nationally representative, includes non-reimbursement costs, and is the only dataset available in Korea that identifies CAM costs.

For our analysis, we cross-sectionally pooled the 4-year data from 2015 to 2018. The reason for pooling was to measure the average use of TKM over the four years rather than to characterize the participants as casual TKM users, as 86 patients with cancer used TKM in 2018, and more than 70% of them used it only once.

The data were accessed between March 15 and April 15, 2024. At no point during or after the data collection process were the researchers provided with any personally identifiable information.

### 2.2. Ethics approval

This study utilized secondary data obtained from Korea Institute for Health and Social Affairs (KIHSA) and National Health Insurance Service (NHIS). Since the research did not involve personally identifiable information, it was exempt from Dongshin University Institutional Review Board (IRB) review (Approval Number: 1040708-202402-SB-045). The study adhered to ethical standards and guidelines as outlined in the Declaration of Helsinki.

### 2.3. Sample selection

The total number of respondents in the 2018 KHPS dataset was 17,008. We defined patients with cancer as those who had cancer in the four years from 2015 to 2018 and selected them as participants if they answered that they had a neoplasm between C00 and D48 in the Korean Standard Classification of Diseases (KCD) regarding their chronic diseases. A total of 358 participants met the criteria, and 321 were selected as the final participants after excluding 37 who did not respond to questions about household income, health insurance type, chronic disease, or quality of life (EuroQoL-5 dimensions; EQ-5D). Of them, 150 (46.72%) were TKM users and 171 (53.27%) were non-users (Figure 1).

**Figure 1.**
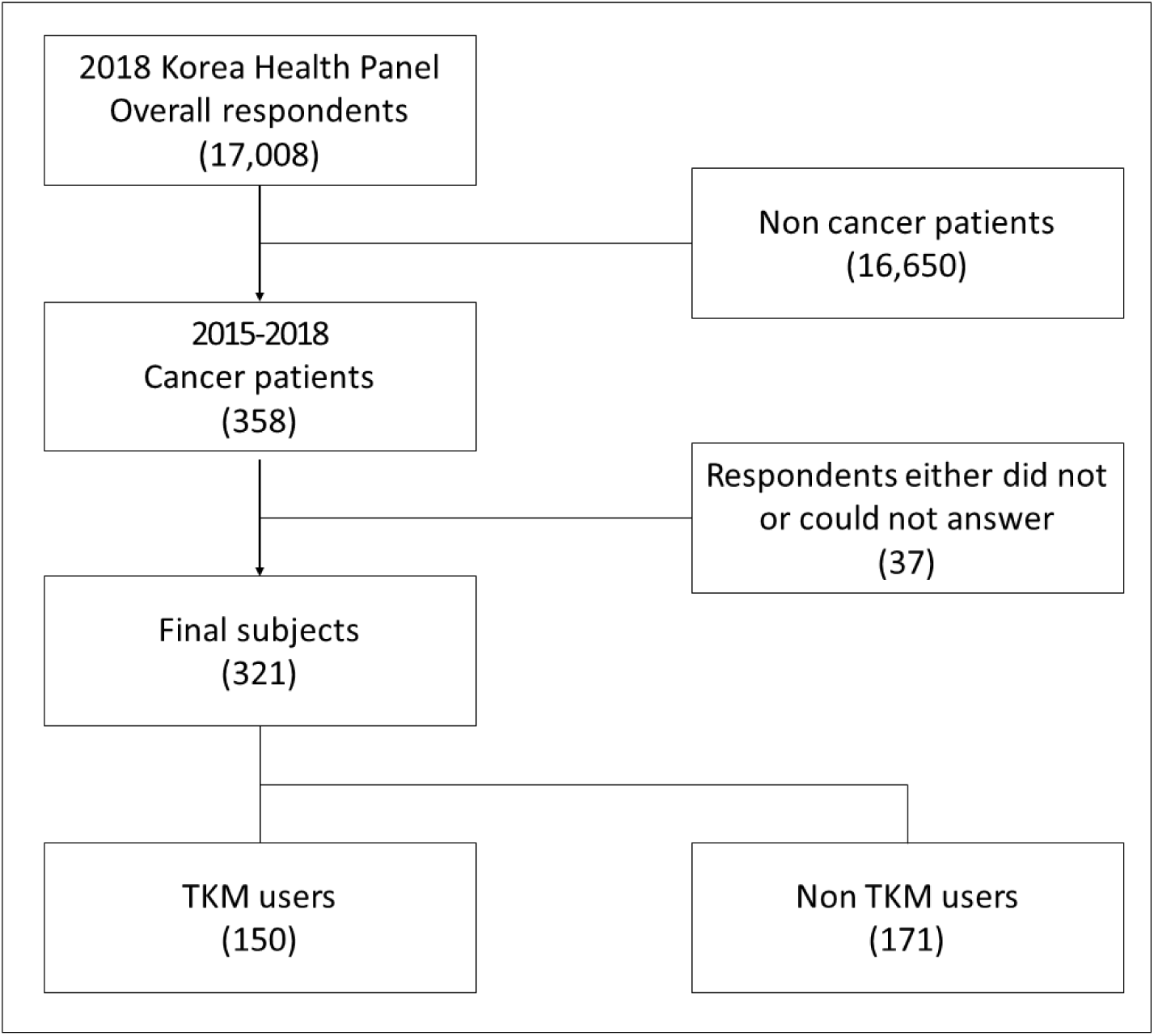
Sample selection. *Notes.* TKM: Traditional Korean Medicine.

### 2.4. Variables

#### 2.4.1. Dependent variables

Two variables related to medical costs and one related to quality of life were analyzed as dependent variables to determine whether they are affected by the patients’ use of TKM. WM cost was selected as the medical cost variable and calculated by summing the total cost (sum of the insurer cost and patient’s out-of-pocket) for all outpatient services, excluding TKM and dentistry, over four years, and averaged annually. Treatment included all treatments for other conditions, not only those for treating cancer. CAM cost was the second cost variable. It referred to the cost of services and products other than medical services used by patients with cancer to improve their health. The KHPS provides information about CAM costs, including cases where acupuncture, herbal medicine, and medicinal herbs are used through pathways other than TKM institutions (Table 1). We combined the CAM costs for the four years and used the annual average.

**Table 1.**
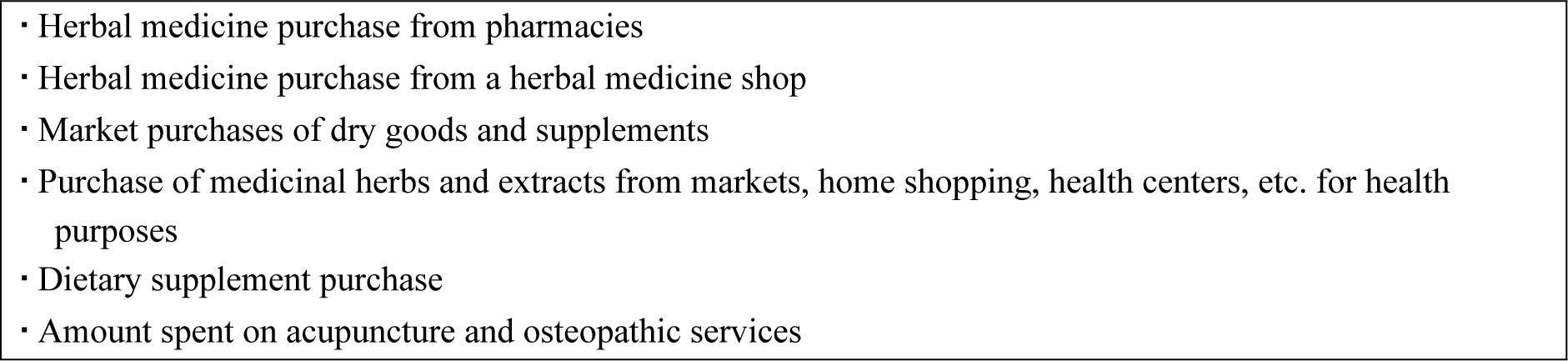
CAM costs from Korea Health Panel Survey (KHPS) Data.

For quality of life variables, we used the EQ-5D. It is a 3-point scale that assesses five domains: exercise capacity, self-care, daily activities, pain/discomfort, and anxiety/depression. The composite index of the EQ-5D was calculated using the South Korean population-based preference [29]. To analyze the impact of TKM use on the change in the quality of life (EQ-5D), the “2018 EQ-5D value – 2015 EQ-5D value” was created to measure the change between 2015 and 2018 and used as the dependent variable (EQ-5D change amount). The EQ-5D was included as a covariate in the model to analyze the impact of using TKM on cost.

#### 2.4.2. Explanatory variables

The explanatory variables used in the study consisted of TKM use and TKM cost. TKM use was defined as the use of TKM outpatient services in the past four years. Participants were categorized as TKM users if they used TKM outpatient services at least once during the four years. The total medical costs (sum of the insurer cost and patient out-of-pocket costs) for all outpatient TKM visits over the four years were summed and averaged annually. Visits for the treatment of all diseases, not only cancer, were included.

#### 2.4.3. Covariates

The covariates were constructed based on the 2018 data and included individual characteristics, healthcare access-related variables, and health status. The individual characteristics included sex, age, and educational level. Educational level was categorized into three groups: elementary school or lower, middle school or higher to high school graduate or lower, and university or higher. The healthcare access-related variable was composed of region, for distance accessibility, and household income, for economic accessibility. The region was classified as “city” if the residence was in one of the 17 metropolitan areas and “town/village” for other regions. To control for the number of household members, the household income was calculated by dividing total household income by the square root of the number of household members. Health-related variables included the number of cancers, presence of three common cancers (thyroid, breast, and stomach cancer), presence of chronic musculoskeletal diseases, Charlson Comorbidity Index (CCI), and quality of life. The number of cancers referred to the type of currently-present cancer and was categorized as one, two, or more. The most common types of cancer were thyroid, breast, and stomach, and their presence or absence were included as variables. Other cancers were rare to be statistically significant and thus, were excluded. In Korea, the most common reason for using TKM is the treatment of musculoskeletal diseases (Health Insurance Review and Assessment Service Medical

Expenses Survey, 2023). Therefore, the presence of chronic musculoskeletal disease was defined as a variable. The CCI, developed by Charlson et al. in 1987, is a measurement tool used to adjust for comorbidities. The validity of converting International Classification of Diseases (ICD)-9 or ICD-10 codes to CCI has been proven in many studies [30], and its predictive power for medical outcomes, such as mortality, has been recognized [31]. The CCI adjusts the sum by assigning weights of 1–6 points to 17 disease groups according to severity. In this study, the participants were assumed to have different needs for using TKM depending on disease severity, and the CCI was reflected as an independent variable to control for this.

EQ-5D was included as an explanatory variable in the model to analyze whether the use of TKM and the cost of TKM treatment affected WM and CAM costs.

### 2.5. Analysis

To analyze the participants’ characteristics, we pooled the data of all years and conducted a descriptive analysis of each variable. For each variable, dependent variables, such as the use and cost of TKM, were analyzed. All continuous variables were analyzed using the t-test or analysis of variance (ANOVA), whereas nominal variables were analyzed using the chi-square test. Next, various behaviors regarding the use of TKM in patients with cancer were analyzed. For behavioral analysis, the rate of TKM use, number of cases of TKM use per person per year, and cost of TKM use per person per year were identified. Additionally, the use of TKM was analyzed by disease or treatment method.

We analyzed the effect of the use of TKM (use and treatment costs) on medical costs and CAM costs in patients with cancer. Prior to the multivariate analysis, a t-test was conducted on the use of TKM, WM costs, and CAM costs to identify the correlation between them. Subsequently, we conducted a generalized linear model (GLM) analysis four times to determine the results after adjusting for participant characteristics. This is because medical cost data were right-skewed and violated the normality assumption of ordinary least squares (OLS). To solve the problem of normality of OLS, it is sometimes transformed into log(y); however, in this case, direct interpretation of the coefficient was difficult, and bias could have been introduced due to retransformation. Therefore, in this study, we used the GLM analysis to determine the data distribution and applied it to the model. The GLM applied the gamma family and log link, which are commonly used in medical cost analyses, and a modified Park test was performed to confirm whether the distribution was appropriate. Additionally, we derived a deviance value to determine the model fit. The closer the deviance value is to 1, the better the model fit. Finally, we presented the average variance inflation factor (VIF) value to identify multicollinearity between the variables. We performed a marginal effect analysis to convert the coefficients into amounts after the analysis. Finally, we performed two regression analyses to examine the effect of TKM use on quality of life (EQ-5D). With quality of life as the dependent variable, two models were analyzed: one with TKM use and the other with TKM cost as the explanatory variable, respectively. After the analysis, the R-square value was presented to demonstrate the explanatory power of the model, and the average VIF was presented to identify multicollinearity among the variables.

## 3. Result

### 3.1. Demographics

Of the 321 participants who reported a cancer diagnosis, 55.50% of women and 32.23% of men used TKM, with a statistically significant difference. The average age of the participants was 66.03 years, and there was no significant age difference between the users and non-users of TKM. By the area of residence, those living in cities spent significantly more on TKM treatment than those living in towns or villages. The average household income of the participants was 23.77±17.98 US$, with non-users having a significantly higher income.

According to the number of diagnosed cancers, those with two or more cancers spent significantly more in medical treatment than those with one. The results showed that the proportion of TKM users was higher among those diagnosed with thyroid cancer.

The utilization rate of TKM was high and TKM costs were significantly higher in the group with chronic musculoskeletal diseases.

Regarding CCI, 196 (61.3%) participants scored 1-2, 87 (27.1%) scored 0, and 37 (11.6%) scored > 3. TKM and CAM costs tended to increase as CCI increased, but did not reach statistical significance, while WM costs tended to increase significantly as CCI increased (Table 2).

**Table 2.** Utilization of TKM and costs of TKM, WM, and CAM according to participant characteristics.

| Variables | Total (N) | Utilization of TKM<br>† (%) | | | TKM costs‡<br>(US\$) | | WM costs‡<br>(US\$) | | CAM costs ‡<br>(US\$) | |
| --- | --- | --- | --- | --- | --- | --- | --- | --- | --- | --- |
|  |  | No | Yes | p-value | Mean±SD | p-value | Mean±SD | p-value | Mean±SD | p-value |
| Total | 321 | 171<br>(53.27) | 150<br>(46.73) |  | 29.71±<br>65.37 |  | 772.25±<br>593.89 |  | 32.09±<br>55.71 |  |
| Sex |  |  |  |  |  |  |  |  |  |  |
| - Men | 121 | 82<br>(67.77) | 39<br>(32.23) | <0.001*** | 18.85±<br>65.37 | 0.059 | 704.45±<br>567.79 | 0.112 | 25.06±<br>35.71 | 0.079 |
| - Women | 200 | 89<br>(44.50) | 111<br>(55.50) |  | 36.29±<br>87.52 |  | 813.26±<br>606.86 |  | 36.34±<br>64.60 |  |
| Age<br>(year,<br>Mean±SD) | 66.03±<br>11.98 | 66.33±<br>12.34 | 65.69±<br>11.59 | 0.637 | - | - | - | - | - | - |
| Educational<br>level |  |  |  |  |  |  |  |  |  |  |
| - Elementary<br>school and<br>below | 92 | 42<br>(45.65) | 50<br>(54.35) | 0.087 | 28.55±<br>64.45 | 0.881 | 835.34±<br>647.53 | 0.482 | 41.20±<br>80.90 | 0.176 |
| - High school<br>and below | 173 | 93<br>(53.76) | 80<br>(46.24) |  | 31.62±<br>87.68 |  | 748.92±<br>570.78 |  | 28.06±<br>40.94 |  |
| - College or<br>higher | 56 | 36<br>(64.29) | 20<br>(35.71) |  | 25.72±<br>80.69 |  | 740.66±<br>574.57 |  | 29.56±<br>42.31 |  |
| Region |  |  |  |  |  |  |  |  |  |  |
| - City | 139 | 68<br>(48.92) | 71<br>(51.08) | 0.172 | 40.24±<br>95.50 | 0.040* | 733.88±<br>574.27 | 0.313 | 32.72±<br>48.79 | 0.859 |
| - Township or<br>village | 182 | 103<br>(56.27) | 150<br>(46.73) |  | 21.67±<br>65.40 |  | 801.55±<br>608.39 |  | 31.61±<br>60.59 |  |
| Household<br>income<br>(1000US\$)§ | 23.77±<br>17.98 | 25.70±<br>19.85 | 21.58±<br>15.34 | 0.040* | - | - | - | - | - | - |
| Number of<br>cancer |  |  |  |  |  |  |  |  |  |  |
| - 1 | 290 | 153<br>(52.76) | 137<br>(47.24) | 0.574 | 31.70±<br>83.16 | 0.176 | 745.35±<br>599.27 | 0.013* | 32.62±<br>57.13 | 0.604 |
| -2 or more | 31 | 18<br>(58.06) | 13<br>(41.94) |  | 11.15±<br>40.62 |  | 1023.88±<br>479.85 |  | 27.15±<br>40.45 |  |
| Thyroid cancer |  |  |  |  |  |  |  |  |  |  |
| - None | 216 | 124<br>(57.41) | 92<br>(42.59) | 0.033* | 25.72±<br>78.38 | 0.201 | 784.31±<br>624.22 | 0.602 | 36.28±<br>64.37 | 0.053 |
| - Yes | 105 | 47<br>(44.76) | 58<br>(55.24) |  | 37.93±<br>83.70 |  | 747.43±<br>528.02 |  | 23.47±<br>29.51 |  |
| Breast cancer |  |  |  |  |  |  |  |  |  |  |
| - None | 284 | 155<br>(54.58) | 129<br>(45.42) | 0.194 | 29.79±<br>78.00 | 0.963 | 749.03±<br>569.29 | 0.052 | 32.59±<br>57.24 | 0.659 |
| - Yes | 37 | 16<br>(43.24) | 21<br>(56.76) |  | 29.13±<br>96.91 |  | 950.46±<br>742.00 |  | 28.28±<br>42.53 |  |
| Stomach cancer |  |  |  |  |  |  |  |  |  |  |
| - None | 268 | 137<br>(51.12) | 131<br>(48.88) | 0.082 | 32.63±<br>83.49 | 0.143 | 794.71±<br>601.08 | 0.128 | 30.70±<br>54.51 | 0.315 |
| - Yes | 53 | 34<br>(64.15) | 19<br>(35.85) |  | 14.96±<br>59.65 |  | 658.66±<br>547.35 |  | 39.13±<br>61.50 |  |
| Chronic<br>musculoskeletal<br>disease |  |  |  |  |  |  |  |  |  |  |
| - None | 144 | 91<br>(63.19) | 53<br>(36.81) | 0.001** | 13.42±<br>52.26 | 0.001** | 680.97±<br>516.17 | 0.013* | 25.92±<br>36.70 | 0.073 |
| - Yes | 177 | 80<br>(45.20) | 97<br>(54.80) |  | 42.97±<br>95.35 |  | 846.51±<br>642.22 |  | 37.11±<br>67.02 |  |
| CCI # |  |  |  |  |  |  |  |  |  |  |
| - 0 | 87 | 49<br>(56.32) | 38<br>(43.68) | 0.798 | 18.09±<br>64.60 | 0.183 | 561.81±<br>484.59 | <0.001*** | 29.46±<br>56.23 | 0.558 |
| - 1-2 | 196 | 102<br>(52.04) | 94<br>(47.96) |  | 31.84±<br>85.18 |  | 825.92±<br>605.33 |  | 31.54±<br>45.64 |  |
| - 3 or more | 38 | 20<br>(52.63) | 18<br>(47.37) |  | 45.31±<br>84.53 |  | 977.20±<br>641.02 |  | 40.94±<br>91.74 |  |
| Quality of life<br>(EQ-5D) | 0.89±<br>0.10 | 0.90±<br>0.10 | 0.89±<br>0.11 | 0.548 | - | - | - | - | - | - |

†: Utilization of TKM over four years (2015–2018) • Results of t-test for continuous variables and chi-square test for nominal variables
‡: Annual average treatment costs for four years (2015-2018) • T-test or analysis of variance (ANOVA) was performed only for nominal variables
§: The adjusted value is calculated by dividing household income by the square root of the number of household members
#: Charlson Comorbidity Index Score
TKM: traditional Korean medicine; CAM: complementary and alternative medicine; WM: Western medicine. \* p<0.05 \*\*p<0.01 \*\*\*p<0.001

### 3.2. Use of TKM by the participants

Of the participants, 86 (26.79%) reported using TKM in 2018. The average number of uses was 2.41±7.22, and the average TKM cost was 26.27±96.04 US$ (Table 3).

**Table 3.**
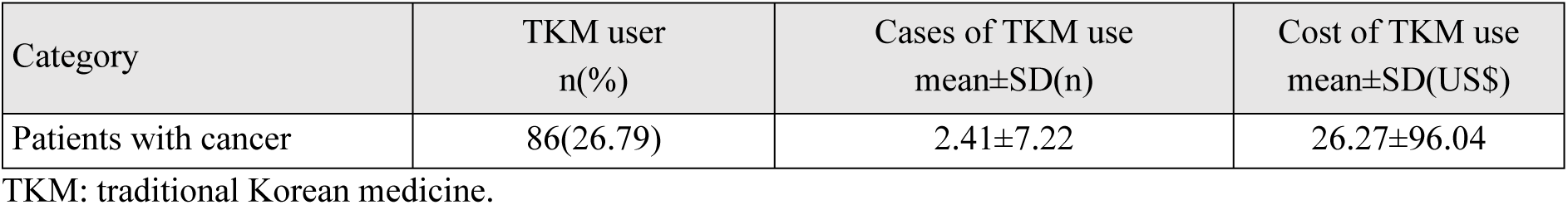
Use of TKM by patients with cancer in 2018 (N=86)

| Category | TKM user<br>n(%) | Cases of TKM use<br>mean±SD(n) | Cost of TKM use<br>mean±SD(US\$) |
| --- | --- | --- | --- |
| Patients with cancer | 86(26.79) | 2.41±7.22 | 26.27±96.04 |

TKM: traditional Korean medicine.

Of the 631 cases that analyzed the disease codes claimed by the National Health Insurance Corporation when using TKM, musculoskeletal and soft tissue diseases were the most frequent with 553 cases (87.64%) and $17,564 (82.74%), followed by treatments with S code (66 cases, 10.46%), a disease code for exogenous damage and injury, with a cost of only $155 (0.73%). The R codes, for abnormal conditions in symptoms and examinations, G codes, for neurological diseases, and others had low rates of less than 5% (Table 4).

**Table 4.** Use of TKM services by diseases in 2018 (N=86)^†^.

| | Cases<br>n(%) | Cost<br>US\$(%) |
| --- | --- | --- |
| Total | 631(100.00) | 21,229(100.00) |
| M00-M99 (Diseases of the musculoskeletal system and connective tissue) | 553(87.64) | 17,564(82.74) |
| S00-T98 (Injury, poisoning and certain other consequences of external cause) | 66(10.46) | 155(0.73) |
| R00-R99 (Symptoms, signs and abnormal clinical and laboratory findings, NEC) | 19(3.01) | 1,604(7.56) |
| G00-G99 (Diseases of the Nervous System) | 15(2.38) | 759(3.58) |
| Others | 13(2.06) | 1,147(5.40) |

†: Duplicate responses
TKM: traditional Korean medicine.

By procedure, acupuncture, moxibustion, and cupping were performed 609 times (96.51%), followed by physical therapy (480 times, 76.07%), and the total treatment costs were $16,264 (76.61%) and $8,491 (40.00%), respectively. There were only 25 herbal medicine prescriptions (3.96%), but the treatment cost was $5,479, accounting for 25.81% of the total treatment cost of TKM (Table 5)..

**Table 5.** Use of TKM services by treatment method in 2018 (N=86)^†^.

| | Cases<br>n(%) | Cost<br>US\$(%) |
| --- | --- | --- |
| Total | 631(100.00) | 21,229(100.00) |
| Acupuncture, moxibustion, and cupping | 609(96.51) | 16,264(76.61) |
| Herbal medicine | 25(3.96) | 5,479(25.81) |
| Chuna | 0(0.00) | 0(0.00) |
| Herbal medicine acupuncture (including Bee-venom Acupuncture) | 13(2.06) | 359(1.69) |
| Physical Therapy | 480(76.07) | 8,491(40.00) |
| Others | 3(0.48) | 22(0.10) |
| <sup>†</sup>: Duplicate responses |  |  |

TKM: traditional Korean medicine.

### 3.3. The impact of the use of TKM on WM and CAM costs

There was no significant difference in WM and CAM costs between users and non-users of TKM (Table 6); even after adjusting for demographic and disease-related variables, TKM cost did not have a significant effect on WM and CAM costs (Table 8). TKM costs did not have a significant correlation with WM costs, but had a weak positive correlation with CAM costs (Table 7). Even after adjusting for demographic and disease variables, there was no significant correlation with WM costs. However, CAM cost increased with TKM cost (Table 9).

**Table 6.** WM and CAM costs according to TKM use.

|  |  | WM cost |  | CAM Cost |  |
| --- | --- | --- | --- | --- | --- |
|  |  | Cost | *p*-value | Cost | *p*-value |
| Use of TKM | No | 739.36±594.42 | 0.290 | 28.69±44.62 | 0.244 |
|  | Yes | 809.73±593.04 |  | 35.96±66.07 |  |

TKM: traditional Korean medicine; CAM: complementary and alternative medicine; WM: Western medicine

**Table 7.** Correlations between TKM, WM, and CAM costs.

|  | WM cost |  | CAM cost |  |
| --- | --- | --- | --- | --- |
|  | Coef | *p*-value | Coef | *p*-value |
| TKM cost | 0.11 | 0.059 | 0.12 | 0.032 |

TKM: traditional Korean medicine; CAM: complementary and alternative medicine; WM: Western medicine

**Table 8.**
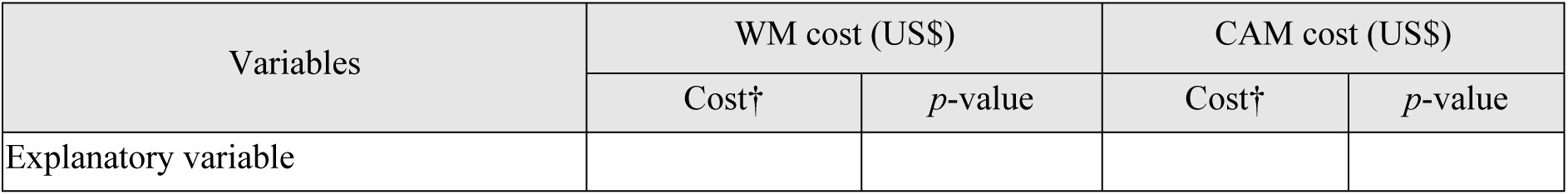
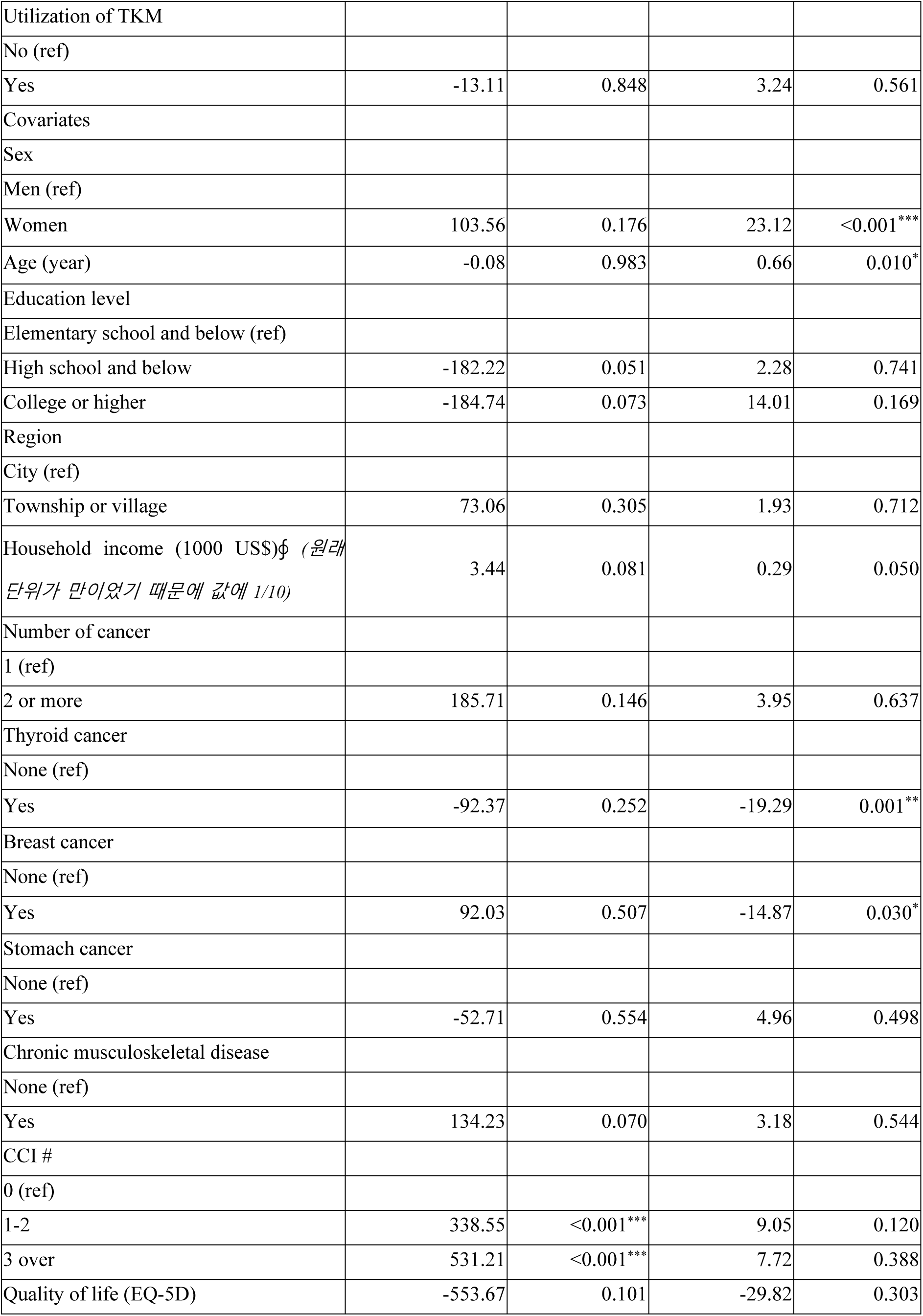
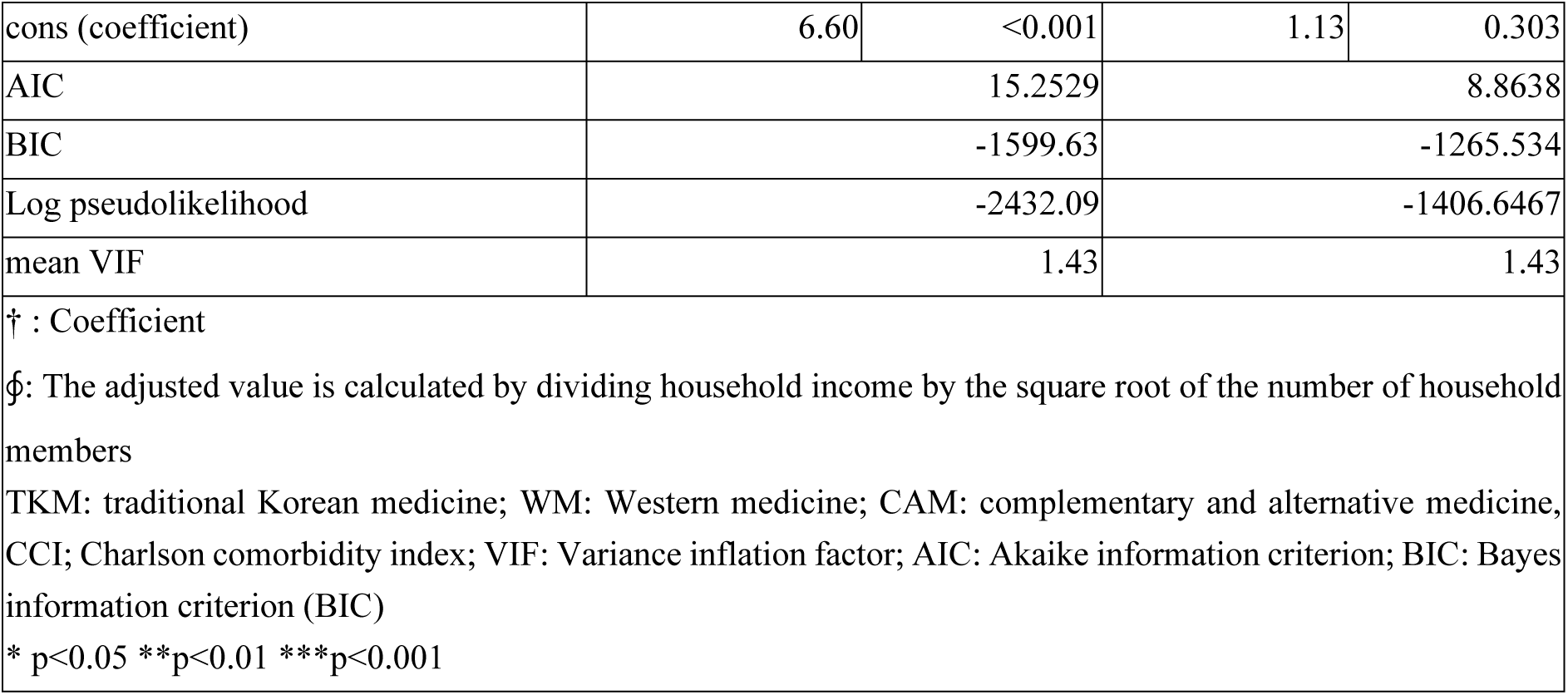
Effect of TKM use on WM and CAM costs.

**Table 9.** Effect of TKM costs on WM and CAM costs.

| Variables | WM cost (US\$) | | CAM cost (US\$) | |
| --- | --- | --- | --- | --- |
|  | Cost† | p-value | Cost† | p-value |
| Explanatory variable |  |  |  |  |
| Cost of TKM | 0.01 | 0.972 | 0.05 | 0.028* |
| Covariates |  |  |  |  |
| Sex |  |  |  |  |
| Men (ref) |  |  |  |  |
| Women | 101.00 | 0.184 | 22.26 | <0.001*** |
| Age (year) | -0.04 | 0.992 | 0.64 | 0.013* |
| Education level |  |  |  |  |
| Elementary school and below (ref) |  |  |  |  |
| High school and below | -180.42 | 0.053 | 2.30 | 0.737 |
| College or higher | -183.71 | 0.074 | 11.45 | 0.237 |
| Region |  |  |  |  |
| City (ref) |  |  |  |  |
| Township or village | 73.64 | 0.305 | 2.34 | 0.649 |
| Household income (1000 US\$)ϕ | 3.44 | 0.082 | 0.27 | 0.059 |
| Number of cancer |  |  |  |  |
| 1 (ref) |  |  |  |  |
| 2 or more | 186.20 | 0.146 | 2.61 | 0.735 |
| Thyroid cancer |  |  |  |  |
| None (ref) |  |  |  |  |
| Yes | -91.37 | 0.260 | -19.13 | 0.001** |
| Breast cancer |  |  |  |  |
| None (ref) |  |  |  |  |
| Yes | 94.18 | 0.498 | -14.91 | 0.025* |
| Stomach cancer |  |  |  |  |
| None (ref) |  |  |  |  |
| Yes | -51.05 | 0.569 | 5.57 | 0.448 |
| Chronic musculoskeletal disease |  |  |  |  |
| None (ref) |  |  |  |  |
| Yes | 132.35 | 0.079 | 1.14 | 0.826 |
| CCI # |  |  |  |  |
| 0 (ref) |  |  |  |  |
| 1-2 | 336.31 | <0.001*** | 8.22 | 0.157 |
| 3 over | 527.36 | <0.001*** | 7.03 | 0.430 |
| Quality of life (EQ-5D) | -553.65 | 0.102 | -25.08 | 0.240 |
| cons | 6.60 | <0.001 | 1.09 | 0.308 |
| AIC | 15.2529 |  | 8.8471 |  |
| BIC | -1599.576 |  | -1270.604 |  |
| Log pseudolikelihood | -2432.0951 |  | -1403.9583 |  |
| mean VIF | 1.43 |  | 1.43 |  |
| † : Coefficient |  |  |  |  |
| ϕ: The adjusted value is calculated by dividing household income by the square root of the number of household members |  |  |  |  |
| TKM: traditional Korean medicine; WM: Western medicine; CAM: complementary and alternative medicine, CCI; Charlson comorbidity index; VIF: Variance inflation factor; AIC: Akaike information criterion; BIC: Bayes information criterion (BIC) |  |  |  |  |
| * p<0.05 **p<0.01 ***p<0.001 |  |  |  |  |

In covariates, the CCI significantly affected WM costs and sex and the diagnosis of thyroid and breast cancers significantly affected CAM costs.

### 3.4. The effect of TKM use on the quality of life

Adjusting for demographic and disease-related variables, the EQ-5D of TKM users increased by 0.03 over four years compared to that of non-users, which was statistically significant. However, the change in EQ-5D values according to the TKM cost was not statistically significant (Table 10).

**Table 10.**
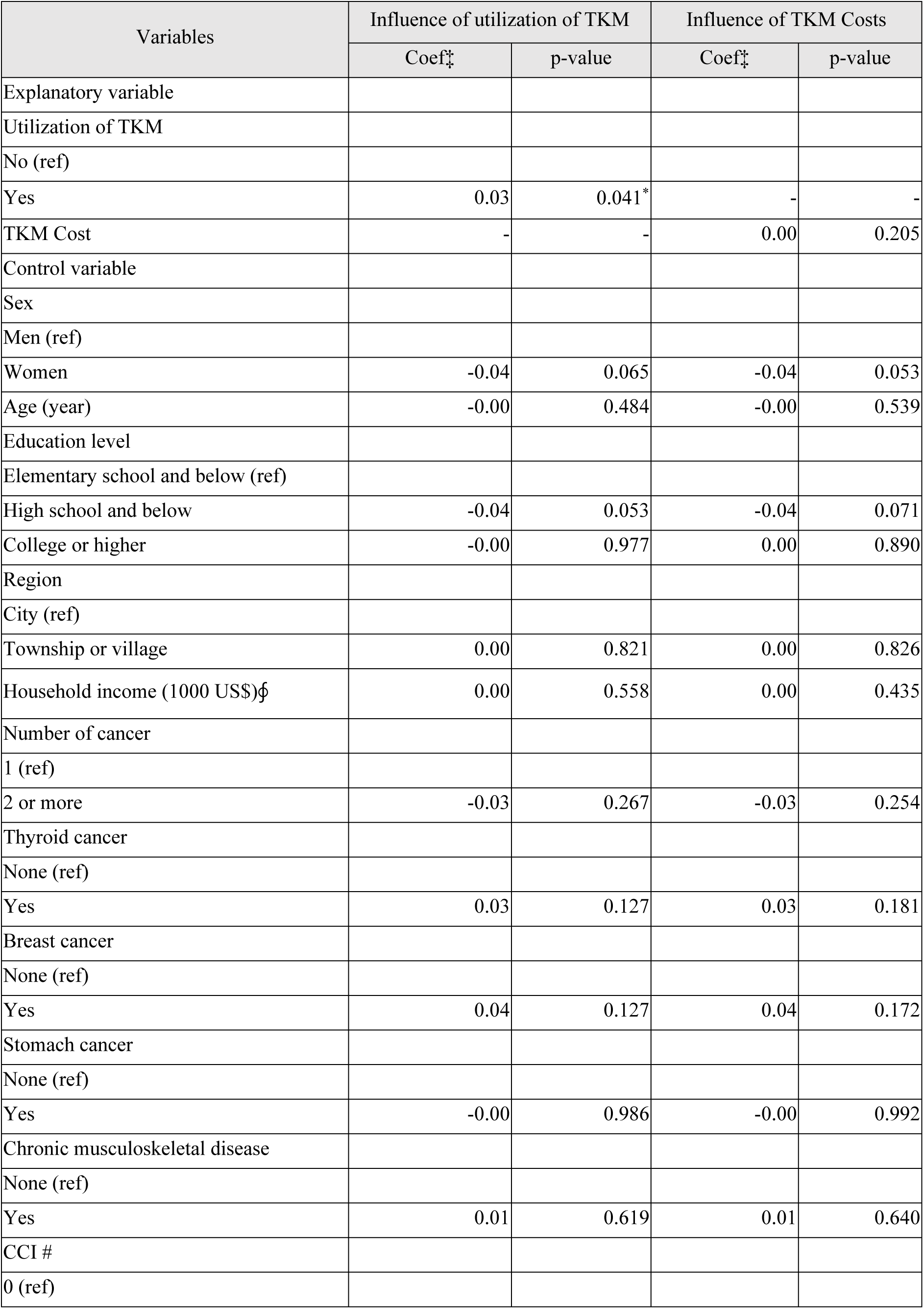
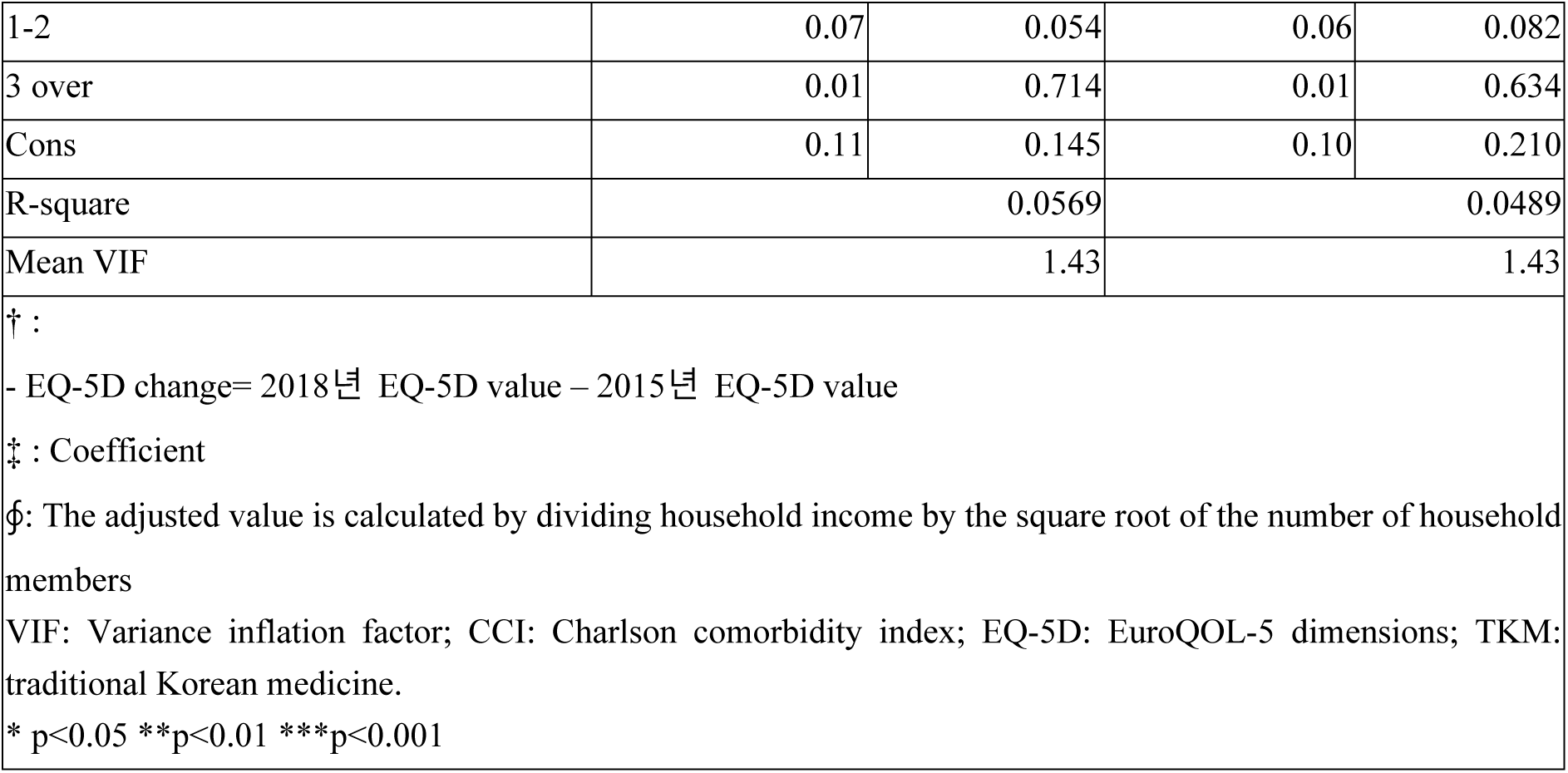
Effect of TKM use on quality of life indicator (EQ-5D)

| Variables | Influence of utilization of TKM |  | Influence of TKM Costs |  |
| --- | --- | --- | --- | --- |
|  | Coef<sup>†</sup> | p-value | Coef<sup>†</sup> | p-value |
| Explanatory variable |  |  |  |  |
| Utilization of TKM |  |  |  |  |
| No (ref) |  |  |  |  |
| Yes | 0.03 | 0.041* | - | - |
| TKM Cost | - | - | 0.00 | 0.205 |
| Control variable |  |  |  |  |
| Sex |  |  |  |  |
| Men (ref) |  |  |  |  |
| Women | -0.04 | 0.065 | -0.04 | 0.053 |
| Age (year) | -0.00 | 0.484 | -0.00 | 0.539 |
| Education level |  |  |  |  |
| Elementary school and below (ref) |  |  |  |  |
| High school and below | -0.04 | 0.053 | -0.04 | 0.071 |
| College or higher | -0.00 | 0.977 | 0.00 | 0.890 |
| Region |  |  |  |  |
| City (ref) |  |  |  |  |
| Township or village | 0.00 | 0.821 | 0.00 | 0.826 |
| Household income (1000 US\$)§ | 0.00 | 0.558 | 0.00 | 0.435 |
| Number of cancer |  |  |  |  |
| 1 (ref) |  |  |  |  |
| 2 or more | -0.03 | 0.267 | -0.03 | 0.254 |
| Thyroid cancer |  |  |  |  |
| None (ref) |  |  |  |  |
| Yes | 0.03 | 0.127 | 0.03 | 0.181 |
| Breast cancer |  |  |  |  |
| None (ref) |  |  |  |  |
| Yes | 0.04 | 0.127 | 0.04 | 0.172 |
| Stomach cancer |  |  |  |  |
| None (ref) |  |  |  |  |
| Yes | -0.00 | 0.986 | -0.00 | 0.992 |
| Chronic musculoskeletal disease |  |  |  |  |
| None (ref) |  |  |  |  |
| Yes | 0.01 | 0.619 | 0.01 | 0.640 |
| CCI # |  |  |  |  |
| 0 (ref) |  |  |  |  |
| 1-2 | 0.07 | 0.054 | 0.06 | 0.082 |
| 3 over | 0.01 | 0.714 | 0.01 | 0.634 |
| Cons | 0.11 | 0.145 | 0.10 | 0.210 |
| R-square | 0.0569 |  | 0.0489 |  |
| Mean VIF | 1.43 |  | 1.43 |  |
| † : |  |  |  |  |
| - EQ-5D change= 2018년 EQ-5D value – 2015년 EQ-5D value |  |  |  |  |
| ‡ : Coefficient |  |  |  |  |
| §: The adjusted value is calculated by dividing household income by the square root of the number of household members |  |  |  |  |
| VIF: Variance inflation factor; CCI: Charlson comorbidity index; EQ-5D: EuroQOL-5 dimensions; TKM: traditional Korean medicine. |  |  |  |  |
| * p<0.05 **p<0.01 ***p<0.001 |  |  |  |  |

## 4. Discussion

This study investigated the effects of TKM use on WM and CAM costs and quality of life in 321 people diagnosed with cancer as per the KMPS. This allowed us to compare TKM use and cost expenditures by demographic characteristics and diseases to confirm which group spent significantly higher. Additionally, through cross-analysis, we could obtain more information on the group of patients with cancer with higher TKM use.

First, in terms of demographic characteristics, female patients with cancer were significantly more likely to be TKM users. This result is consistent with those of previous studies showing that women are more likely to use CAM [32]. In terms of household income, unlike overseas studies [33], which generally reported a higher proportion of CAM users in the middle class or higher, this study found that the average household income of TKM users is lower than that of non-users. However, there are a few reports [34] such as the results of the aging panel survey conducted in Korea, which reported that the utilization rate of TKM among those with low income was high, consistent with the findings of this study. This is probably because TKM is covered by the NHIS, making it relatively easily accessible to individuals with low income. In addition, if users are professionals or with high incomes, delays in receiving medical services due to work-related issues may have an impact [35].

According to disease characteristics, the findings showed that when the CCI was high, TKM or CAM costs did not increase significantly, whereas WM costs did. In the calculation formula for CCI [36], a value of 0 implies that cancer is not present currently; therefore, it corresponds to the case of survivors who have been diagnosed with cancer. A case of CCI 1 or higher corresponds to patients diagnosed with cancer or other system diseases that significantly affect life expectancy, such as cardiovascular, respiratory, or neurological diseases. In other words, compared with patients who have completely recovered, those who are currently being treated for cancer or other diseases spend more on medical costs. In the case of chronic musculoskeletal diseases, the number of TKM users and the expenditure were high; however, as the presence of musculoskeletal diseases did not affect CCI, a weak correlation was found between TKM costs and CCI.

Next, we confirmed whether TKM use and costs significantly affected expenditures for WM and CAM and attempted to indirectly infer the current position of TKM in terms of medical economics in patients with cancer.

TKM costs did not correlate with WM costs, and GLM through analysis of covariance also showed that the use of TKM and costs did not affect WM costs. In general, when inferring the relationship between CAM and biomedicine, a positive correlation in cost can be interpreted as a complementary relationship, whereas a negative correlation can be interpreted as a substitute relationship. Accordingly, the results of this study suggest that the pattern of utilization of TKM and WM costs do not have a complementary/substitute relationship but are independent of each other.

TKM costs positively correlated with CAM costs. Considering that TKM use did not have a significant relationship with CAM costs, expenditures in specific areas that increase TKM costs may be positively correlated with CAM costs. For example, considering the use of TKM, herbal medicine was used in only about 3% of treatment cases, but accounted for approximately 25% of the total TKM costs. Most TKM users who use low-cost treatments, such as acupuncture and physical therapy, do not spend in CAM; however, if a small number of patients who are prescribed herbal medicines invest in CAM costs, a model independent of use but correlated with costs can be derived. This means that consumers who receive herbal medicine prescriptions in TKM institutions and those who purchase health-functional foods and herbal medicines from pharmacies, herbal medicine shops, etc., are similarly formed and are not mutually substituted.

Finally, we conducted a regression analysis to determine the effects of TKM use and cost on quality of life indicators, measuring the degree of improvement in the quality of life of patients with cancer based solely on the use of TKM, excluding the influence of other factors.

Regression analysis of the effects of TKM use on the quality of life indicator (EQ-5D) showed that TKM users had a higher quality of life than non-users. A previous study comparing patients who received TWM combination treatment and conventional treatment for breast cancer showed that the TWM group recorded a significantly higher score in EuroQOL visual analogue scale (EQ-VAS) than the non-TWM group and found that European Organization for Research and Treatment of Cancer (EORTC) Core Quality of Life Questionnaire (QLC-30), physical, emotional, cognitive, and function were significantly related to the improvement in quality of life [37]. In addition, acupuncture, which was most frequent in the participants in this study, can improve the quality of life by improving the Karnofsky Performance Score of patients with cancer who are terminally ill [38]. Considering that the indicators used in the quality of life index [39] are mobility, self-care, usual activities, pain/discomfort, and anxiety/depression, those who used TKM received additional measures to improve their quality of life, and the use could improve areas that could not be resolved with existing biomedicine. This suggests that there is a possibility of a positive effect when using TKM and WM together, which means that they may also affect the overall medical costs for patients with cancer.

To the best of our knowledge, this is the first study to analyze the correlation between TKM use, cost, and quality of life in patients diagnosed with cancer using the Korean Medical Panel Data. However, considering the form of the study and the characteristics of the extracted samples, the following limitations exist.

First, there may have been attrition bias due to the characteristics of the medical panel. Patients with cancer who were physically or mentally incapable of participating in the medical panel survey were excluded from the panel selection. Accordingly, the panel data may not have included a population with moderate or higher cancer-related pain.

Second, the characteristics of “TKM users” were selected broadly; therefore, many participants who only used TKM in areas not directly related to cancer may have been included. When the claim ratios of M and S codes were added, more than 85% of the total number of treatments were recorded, and the codes corresponding to neoplasms and tumors were insufficient to be counted separately. In Korea, a significant proportion of TKM use is associated with musculoskeletal diseases [40]. Considering that the rate of TKM use in patients with chronic musculoskeletal diseases was significantly higher in this study, it is presumed that this factor was also included in the number of patients with cancer who used TKM to treat musculoskeletal diseases. Considering this, the lack of correlation between TKM use and WM costs mentioned in the previous review may have been due to dispersion in the purpose of medical care use. In other words, WM costs were probably spent on managing cancer and internal diseases, and TKM costs were mostly used to treat musculoskeletal diseases, injuries, etc. Therefore, this should be interpreted by considering the possibility that the correlation between the two variables was diluted.

Accordingly, in the future, it will be necessary to study the TKM usage behavior of patients with cancer through qualitative research. To investigate the cost of TKM for cancer treatment, it is necessary to utilize all health insurance data by limiting them to codes related to tumors and neoplasms. However, since this also generates information limited to reimbursement items, it may be difficult to investigate non-reimbursed items; nevertheless, it will be meaningful to investigate the cost expenditures for the parts currently covered through reimbursement.

Finally, it may be possible to estimate costs through a registry study of patients who visit TKM institutions and compare them with those of patients who only visit WM institutions. It could be a more valuable study because of the estimation of all covered and non-covered items, including quality of life and other indicators that can be assessed through surveys.

## 5. Conclusions

This study found that the costs of TKM use by patients with cancer were positively correlated with CAM costs. However, it was not due to the use of TKM. It can be inferred that the increase in specific costs, not identified in this study, affected TKM and CAM costs. However, the use of TKM and its costs were not significantly related to the WM costs. This suggests that WM and TKM have independent relationships rather than complementary or alternative relationships for patients with cancer. However, it may be because the purpose of TKM use in this study was not limited to cancer treatment. In addition, the quality of life improved in TKM users, which implies that TKM addressed the problems that could not be solved by WM. This suggests that the appropriate use of WM and TKM may improve cancer-related symptoms and quality of life and control overall medical costs.

## Data Availability

All relevant data are within the manuscript and its Supporting Information files.

## List of abbreviations

TKM: Traditional Korean medicine
TM: traditional medicine
CAM: complementary and alternative medicine
NHIS: National Health Insurance Service
KHPS: Korea Health Panel Survey
KIHASA: Korea Institute for Health and Social Affairs.

